# Should Coronary Revascularization Precede Transcatheter Aortic Valve Replacement? A Meta-Analysis of Randomized Controlled Trials

**DOI:** 10.64898/2026.05.15.26353318

**Authors:** Dina Soliman, John Abdelmalek, Chanokporn Puchongmart, Tulaton Sodsri, Nithila Sivakumar, Zhaunn Sly

## Abstract

**Background:** In severe aortic stenosis patients undergoing TAVR, whether coexisting coronary disease prompts revascularization and its optimal timing remain unclear.

**Aim:** To evaluate the efficacy and safety of PCI before TAVR compared to deferred PCI in patients with severe aortic stenosis and concomitant coronary artery disease.

**Methods:** We performed a meta-analysis of RCTs. PubMed, Embase, Scopus, CENTRAL, and Web of Science were searched for RCTs comparing PCI before TAVR versus no PCI. HRs with 95% CIs were pooled using random-effects models.

**Results:** Three RCTs (ACTIVATION, NOTION-3, PRO-TAVI) enrolling 1,156 patients (579 PCI, 577 no-PCI) were included. Routine PCI before TAVR did not reduce all-cause mortality (HR 0.88, 95% CI 0.67–1.17; p=0.38) or cardiovascular death (HR 0.77, 95% CI 0.49–1.19; p=0.23). PCI significantly reduced any revascularization (HR 0.24, 95% CI 0.06–0.86; p=0.029), and urgent revascularization (HR 0.33, 95% CI 0.12–0.87; p=0.025). MI was not significantly reduced with PCI (HR 0.84, 95% CI 0.44–1.59; p = 0.59). Stroke showed a borderline trend favoring PCI (HR 0.69, 95% CI 0.46–1.04; p=0.073). PCI significantly increased any bleeding (HR 1.96, 95% CI 1.28–3.0; p=0.002) and major bleeding (HR 1.88, 95% CI 1.07–3.31, p=0.027). Neither AKI nor rehospitalization differed significantly between groups. Leave-one-out sensitivity analyses confirmed the stability of mortality, stroke, and bleeding estimates.

**Conclusions:** Routine PCI before TAVR does not reduce mortality. It lowers urgent revascularization and trends toward less stroke but nearly doubles bleeding. Findings support selective, individualized PCI rather than routine revascularization before TAVR.

## 1. INTRODUCTION

Transcatheter aortic valve replacement (TAVR) has evolved from a therapy for inoperable or high-risk patients to the standard of care for older patients with symptomatic severe aortic stenosis (AS), regardless of surgical risk [1]. Concomitant CAD is reported in 40–75% of patients undergoing TAVR [2].

In stable CAD without valvular disease, landmark trials such as ISCHEMIA and COURAGE, along with multiple meta-analyses, have shown that elective revascularization confers no survival advantage over optimal medical therapy [3, 4]. However, the pathophysiology of myocardial ischemia in severe AS may differ fundamentally from that of isolated CAD. AS induces left ventricular hypertrophy, increasing myocardial oxygen demand while compromising supply through capillary rarefaction, perivascular fibrosis, reduced diastolic perfusion time, and impaired coronary flow reserve [5]. This supply–demand mismatch renders the myocardium—particularly the subendocardium—vulnerable to ischemia even without epicardial obstruction [6]. Symptoms are also difficult to attribute solely to AS or CAD. Physiologic assessment differs as well: Minten et al. proposed FFR ≤0.83 and RFR ≤0.85 in AS, compared with ≤0.8 and ≤0.89 in isolated CAD [7].

Despite this rationale, percutaneous coronary intervention (PCI) carries procedural risks, including vascular complications, periprocedural MI, and contrast-induced nephropathy. PCI also necessitates dual antiplatelet therapy (DAPT), consistently linked to increased bleeding in TAVR populations [8]—a particular concern given the elderly demographic with high comorbidity burden predisposing to hemorrhagic events.

Current U.S. and European guidelines do not clearly recommend for or against routine PCI in TAVR patients with concomitant CAD. Existing evidence is conflicting: some randomized trials suggest benefit, others show none, and observational studies and meta-analyses yield heterogeneous findings. Given this uncertainty, we conducted a systematic review and meta-analysis to evaluate the impact of PCI on clinical outcomes in patients with concomitant CAD undergoing TAVR, aiming to provide a more definitive answer to this clinically important question.

## 2. METHODS

### 2.1 Study Registration and Protocol

This systematic review and meta-analysis were conducted in accordance with the Preferred Reporting Items for Systematic Reviews and Meta-Analyses (PRISMA) 2020 guidelines. The protocol was prospectively registered with PROSPERO [CRD420261364189].

### 2.2 Eligibility Criteria

Eligible studies were randomized controlled trials comparing PCI before TAVR (intervention) with conservative management, no PCI, or deferred PCI strategy (comparator) in adult patients (≥18 years) with severe symptomatic aortic stenosis scheduled for TAVR and concomitant coronary artery disease. Non-randomized studies, observational studies, and trials comparing PCI after TAVR versus before TAVR (sequencing studies) or those in patients undergoing surgical aortic valve replacement were excluded.

### 2.3 Information Sources and Search Strategy

We searched PubMed, Embase, Scopus, Cochrane Central Register of Controlled Trials (CENTRAL), and Web of Science using Medical Subject Headings (MeSH) terms and free-text keywords related to transcatheter aortic valve replacement (aortic stenosis, TAVR, TAVI, transcatheter aortic valve implantation/replacement) and percutaneous coronary intervention (PCI, coronary angioplasty, coronary revascularization). The search was limited to randomized controlled trials published in English between 2010 and April 2026. Records were imported into Covidence for screening and data management.

### 2.4 Study Selection and Data Extraction

Two reviewers independently screened titles, abstracts, and full texts, with discrepancies resolved by a third reviewer. Data were extracted independently using a standardized Excel sheet. Corresponding authors were contacted when necessary to obtain missing data.

### 2.5 Outcomes

The primary outcome was all-cause mortality. Secondary outcomes included cardiovascular mortality, myocardial infarction, stroke, any bleeding, major bleeding, any revascularization, urgent revascularization, acute kidney injury, and rehospitalization.

### 2.6 Risk of Bias Assessment

Risk of bias was assessed using the Cochrane Risk of Bias 2 (RoB-2) tool for randomized trials. Each trial was evaluated across five domains: randomization process, deviations from intended interventions, missing outcome data, measurement of outcomes, and selection of reported results. Two reviewers independently assessed each domain, with disagreements resolved by discussion.

### 2.7 Statistical Analysis

The principal effect measure was the hazard ratio (HR), prioritized over crude risk ratios to account for differing follow-up durations. We contacted NOTION-3 investigators for 1-year event-level data; they acknowledged the request but could not share data due to an ongoing separate analysis.

The primary analysis pooled HRs using inverse-variance weighting under a random-effects model (DerSimonian–Laird estimator of between-study variance [τ²]). Reported HRs and corresponding 95% confidence intervals (CIs) were extracted, log-transformed internally, and exponentiated for presentation.

Pooled baseline characteristics by treatment arm were compared using the chi-squared test with Yates correction for categorical variables and Welch’s two-sample t-test for continuous variables. For trials reporting median (IQR), standard deviation was approximated as IQR/1.35; STS and SYNTAX scores were not formally tested due to skewed distributions.

Statistical heterogeneity was assessed using the I² statistic. Random-effects models were applied regardless of heterogeneity.

Leave-one-out sensitivity analyses sequentially excluded each study to evaluate the robustness of pooled estimates. All analyses were performed in R (R Foundation, Vienna, Austria) with the meta package. Two-sided p < 0.05 was considered significant.

## 3. RESULTS

### 3.1 Study Selection

We initially identified 672 studies through database searches. After removing duplicates and ineligible records, 341 studies were screened, and 11 full-text articles were reviewed in detail.

Ultimately, 3 randomized controlled trials met the inclusion criteria and were included in the final analysis; ACTIVATION[9], NOTION-3 [10], and PRO-TAVI [11] **(Figure 1)**.

**Figure 1.**
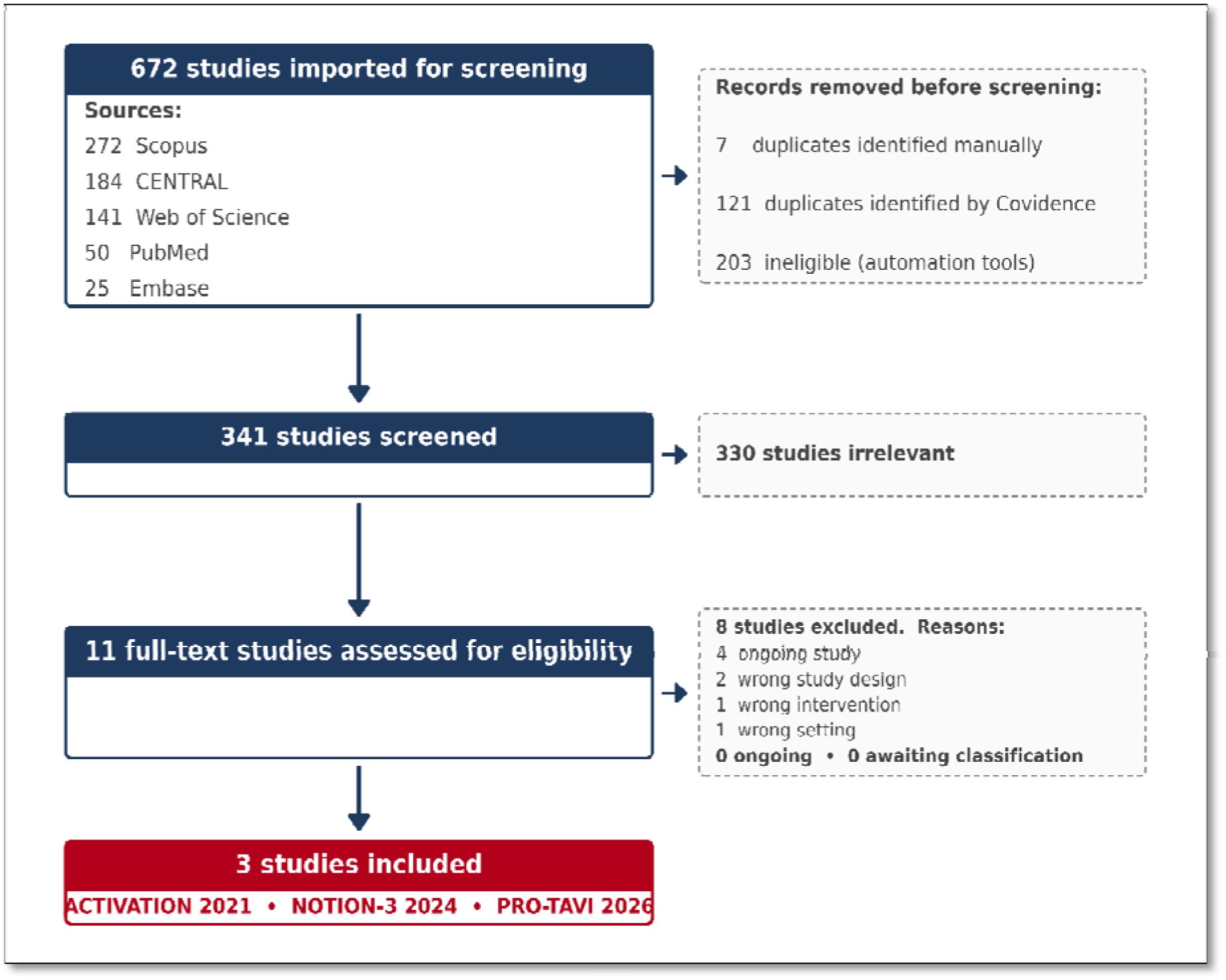
PRISMA Flow Diagram of Study Selection Process. This figure illustrates the systematic literature search and screening process following PRISMA guidelines. A total of 672 records were identified across 5 databases (Scopus, CENTRAL, Web of Science, PubMed, Embase). After removing 128 duplicates and 203 ineligible records via automation tools, 341 studies were screened. Of these, 330 were deemed irrelevant, leaving 11 full-text articles assessed for eligibility. Eight were excluded (4 ongoing, 2 wrong study design, 1 wrong intervention, 1 wrong setting), yielding 3 included RCTs: ACTIVATION (2021), NOTION-3 (2024), and PRO-TAVI (2026). Downward arrows (↓) indicate progression through screening stages; rightward arrows (→) indicate excluded records at each stage. Abbreviations: CENTRAL, Cochrane Central Register of Controlled Trials; PRISMA, Preferred Reporting Items for Systematic Reviews and Meta-Analyses; RCT, randomized controlled trial.

### 3.2 Study Characteristics

The three trials enrolled 1,156 patients (579 PCI, 577 no PCI). ACTIVATION was conducted across 17 centers in the UK, France, and Germany; NOTION-3 across the Nordic-Baltic region; and PRO-TAVI at 12 Dutch hospitals. All were multicenter and open-label. ACTIVATION was prematurely terminated at 235 of the planned 310 patients **(Table 1).**

**Table 1.**
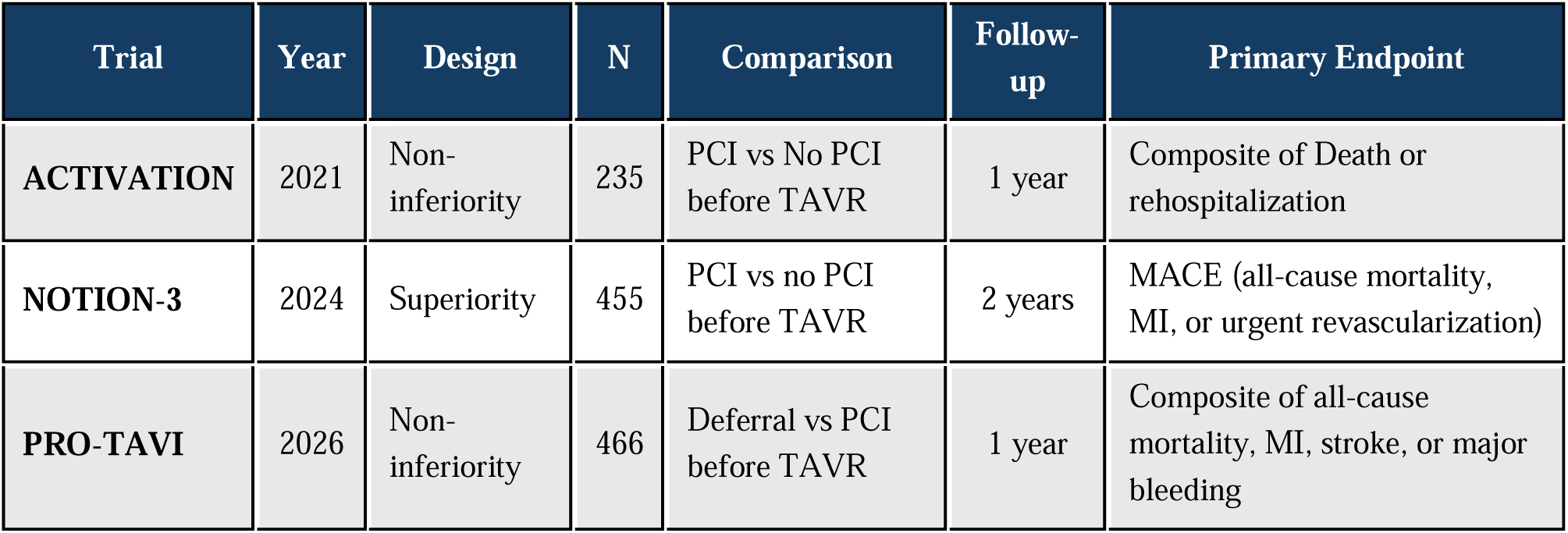
Three randomized controlled trials included.

### 3.3 Patient Characteristics

Pooled baseline characteristics of the 1,156 patients included in the meta-analysis are summarized in **Table 2**. The cohort consisted of elderly patients, of whom approximately two-thirds were male and half of them were symptomatic, with NYHA functional class III–IV. Cardiovascular risk factors were prevalent, including hypertension, hypercholesterolemia, and diabetes mellitus. Prior coronary intervention, CABG, and MI were relatively uncommon, suggesting that coronary artery disease in this population was largely identified during the TAVR work-up rather than reflecting longstanding established disease. Coronary anatomy was predominantly of low complexity, with a median SYNTAX score of 9–10 across the trials reporting this variable, and surgical risk was low to intermediate (median STS score < 5%) **(Table 2)**.

**Table 2.**
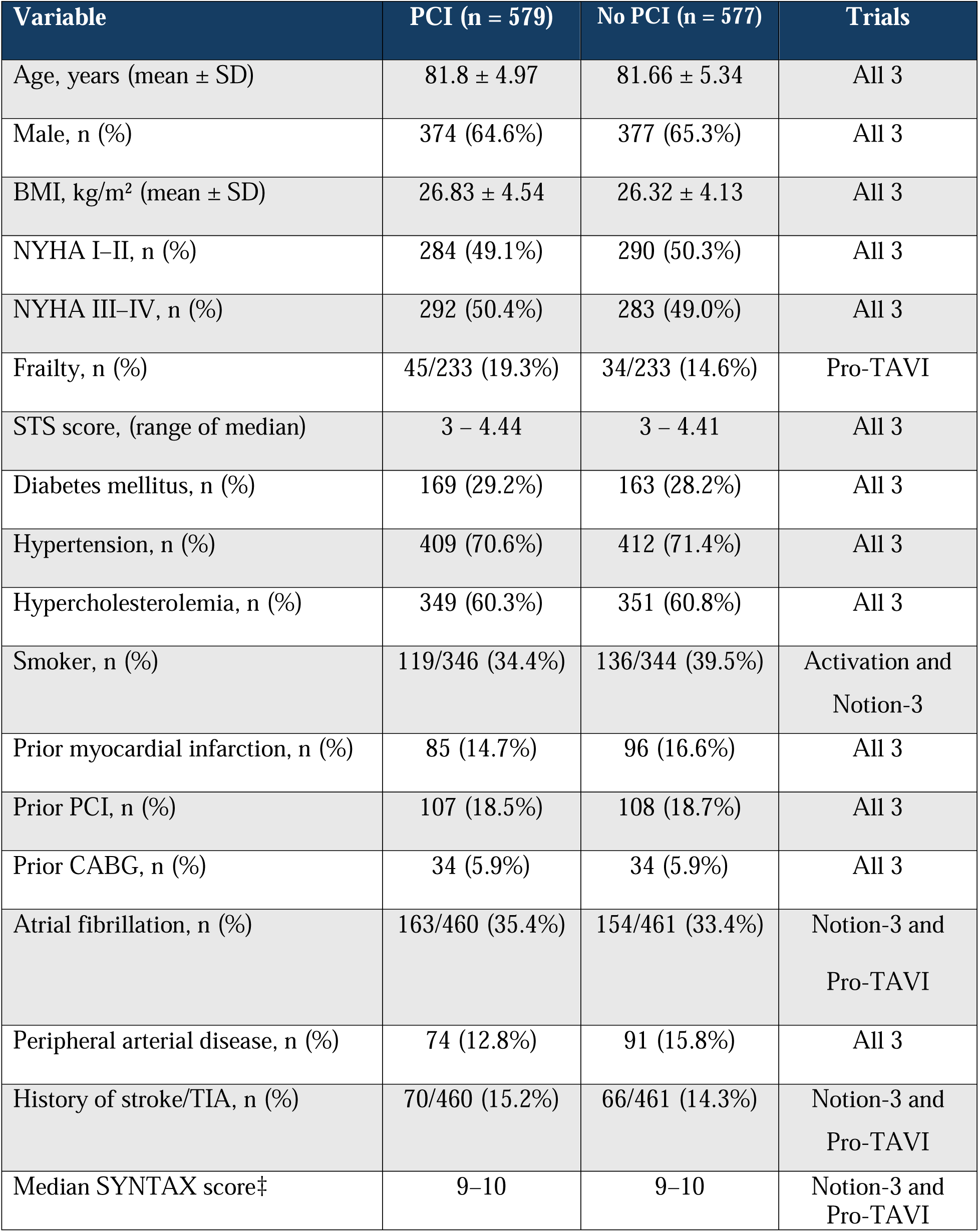
Pooled Patient Characteristics of the Included Randomized Controlled Trials.

| Variable | PCI (n = 579) | No PCI (n = 577) | Trials |
| --- | --- | --- | --- |
| Age, years (mean ± SD) | 81.8 ± 4.97 | 81.66 ± 5.34 | All 3 |
| Male, n (%) | 374 (64.6%) | 377 (65.3%) | All 3 |
| BMI, kg/m<sup>2</sup> (mean ± SD) | 26.83 ± 4.54 | 26.32 ± 4.13 | All 3 |
| NYHA I–II, n (%) | 284 (49.1%) | 290 (50.3%) | All 3 |
| NYHA III–IV, n (%) | 292 (50.4%) | 283 (49.0%) | All 3 |
| Frailty, n (%) | 45/233 (19.3%) | 34/233 (14.6%) | Pro-TAVI |
| STS score, (range of median) | 3 – 4.44 | 3 – 4.41 | All 3 |
| Diabetes mellitus, n (%) | 169 (29.2%) | 163 (28.2%) | All 3 |
| Hypertension, n (%) | 409 (70.6%) | 412 (71.4%) | All 3 |
| Hypercholesterolemia, n (%) | 349 (60.3%) | 351 (60.8%) | All 3 |
| Smoker, n (%) | 119/346 (34.4%) | 136/344 (39.5%) | Activation and<br>Notion-3 |
| Prior myocardial infarction, n (%) | 85 (14.7%) | 96 (16.6%) | All 3 |
| Prior PCI, n (%) | 107 (18.5%) | 108 (18.7%) | All 3 |
| Prior CABG, n (%) | 34 (5.9%) | 34 (5.9%) | All 3 |
| Atrial fibrillation, n (%) | 163/460 (35.4%) | 154/461 (33.4%) | Notion-3 and<br>Pro-TAVI |
| Peripheral arterial disease, n (%) | 74 (12.8%) | 91 (15.8%) | All 3 |
| History of stroke/TIA, n (%) | 70/460 (15.2%) | 66/461 (14.3%) | Notion-3 and<br>Pro-TAVI |
| Median SYNTAX score‡ | 9–10 | 9–10 | Notion-3 and<br>Pro-TAVI |

### 3.4 Risk of Bias

Risk of bias was assessed for each included RCT using the Cochrane Risk of Bias 2 (ROB-2) tool across all five domains.

ACTIVATION was judged at overall some concerns, primarily due to premature termination (235 of 310 planned patients), incomplete follow-up (94.2%), and unblinded clinical events committee, potentially influencing subjective endpoint adjudication. The trial also reflected an earlier era of TAVR practice (2012–2019) with higher surgical risk scores and bare-metal stents in ∼20% of PCI patients, limiting generalizability.

NOTION-3 was judged at overall some concerns. Although randomization was adequate and endpoint adjudication was blinded, the open-label design, protocol-permitted PCI timing flexibility (26% performed concomitantly with or after TAVR), and mid-trial changes to antithrombotic regimens following POPular TAVI and AUGUSTUS introduced co-intervention heterogeneity.

PRO-TAVI was judged at overall some concerns. The primary composite combined directionally opposing outcomes—ischemic events and major bleeding—which move oppositely with PCI, and the 11-percentage-point non-inferiority margin lacks empirical justification. Nonetheless, operational quality was strong: adequate concealment, near-complete follow-up (>99%), and blinded independent endpoint review using VARC-3 definitions.

No trial was judged at high risk of bias, and no selective outcome reporting was identified. Funnel plots and Egger’s test were not performed given few included studies.

### 3.5 Primary Outcome: All-Cause Mortality

All-cause mortality, reported in all three trials (n = 1,156), did not differ significantly between groups (HR 0.88, 95% CI 0.67–1.17; p = 0.378), with no observed heterogeneity (I² = 0.0%, τ² = 0.000) **(Figure 2A)**.

**Figure 2.**
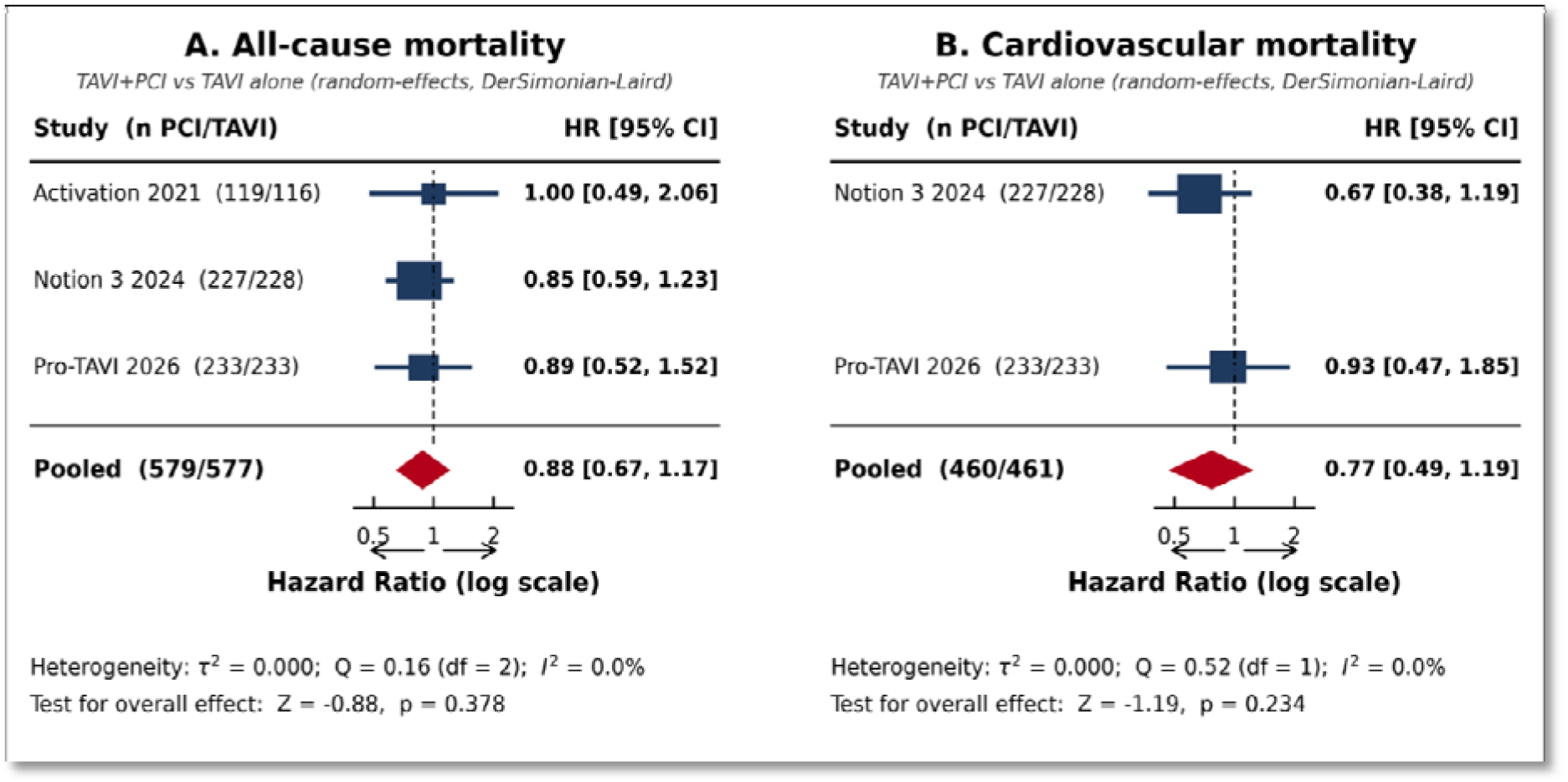
Forest Plots of Mortality Outcomes: PCI Plus TAVR Versus TAVR Alone. This figure presents random-effects (DerSimonian-Laird) meta-analyses of mortality endpoints across three RCTs. Panel A shows all-cause mortality (pooled HR 0.88, 95% CI 0.67–1.17; p=0.378; I²=0.0%), and Panel B shows cardiovascular mortality (pooled HR 0.77, 95% CI 0.49–1.19; p=0.234; I²=0.0%). Neither endpoint reached statistical significance, indicating routine PCI before TAVR does not reduce mortality. Blue squares represent individual trial HRs, with square size proportional to study weight; horizontal lines denote 95% CIs. Red diamonds represent pooled estimates. The dashed vertical line marks HR=1.0 (no effect). Abbreviations: CI, confidence interval; df, degrees of freedom; HR, hazard ratio; I², I-squared statistic for heterogeneity; n, number of patients; PCI, percutaneous coronary intervention; Q, Cochran’s Q statistic; TAVI, transcatheter aortic valve implantation; τ², tau-squared; Z, test statistic for the overall effect.

### 3.6 Cardiovascular Death

Cardiovascular mortality was reported in NOTION-3 and Pro-TAVI. NOTION-3 reported 20 events in the PCI arm vs. 30 in the no-PCI arm (HR 0.67, 95% CI 0.38-1.19). Pro-TAVI reported 16 vs. 17 events (HR 0.93, 95% CI 0.47-1.85). The pooled estimate showed no significant difference, with a numerical trend favoring TAVR plus PCI (HR 0.77, 95% CI 0.49–1.19; p = 0.234; I² = 0.0%). CV mortality HR was not available for Activation 2021 and thus that trial was excluded from this pooled estimate **(Figure 2B).**

### 3.7 Revascularization outcomes

TAVR plus PCI was associated with marked reductions in subsequent revascularization. The risk of any revascularization, reported by NOTION-3 and Pro-TAVI, was substantially lower in the combined-procedure arm (pooled HR 0.24, 95% CI 0.06–0.86; p = 0.029); however, between-trial heterogeneity was high (I² = 81.7%, τ² = 0.713), reflecting a considerably larger effect size in NOTION-3 than in Pro-TAVI. Urgent revascularization was also significantly reduced (pooled HR 0.33, 95% CI 0.12–0.87; p = 0.025; I² = 54.5%) **(Figure 3A, 3B).**

**Figure 3.**
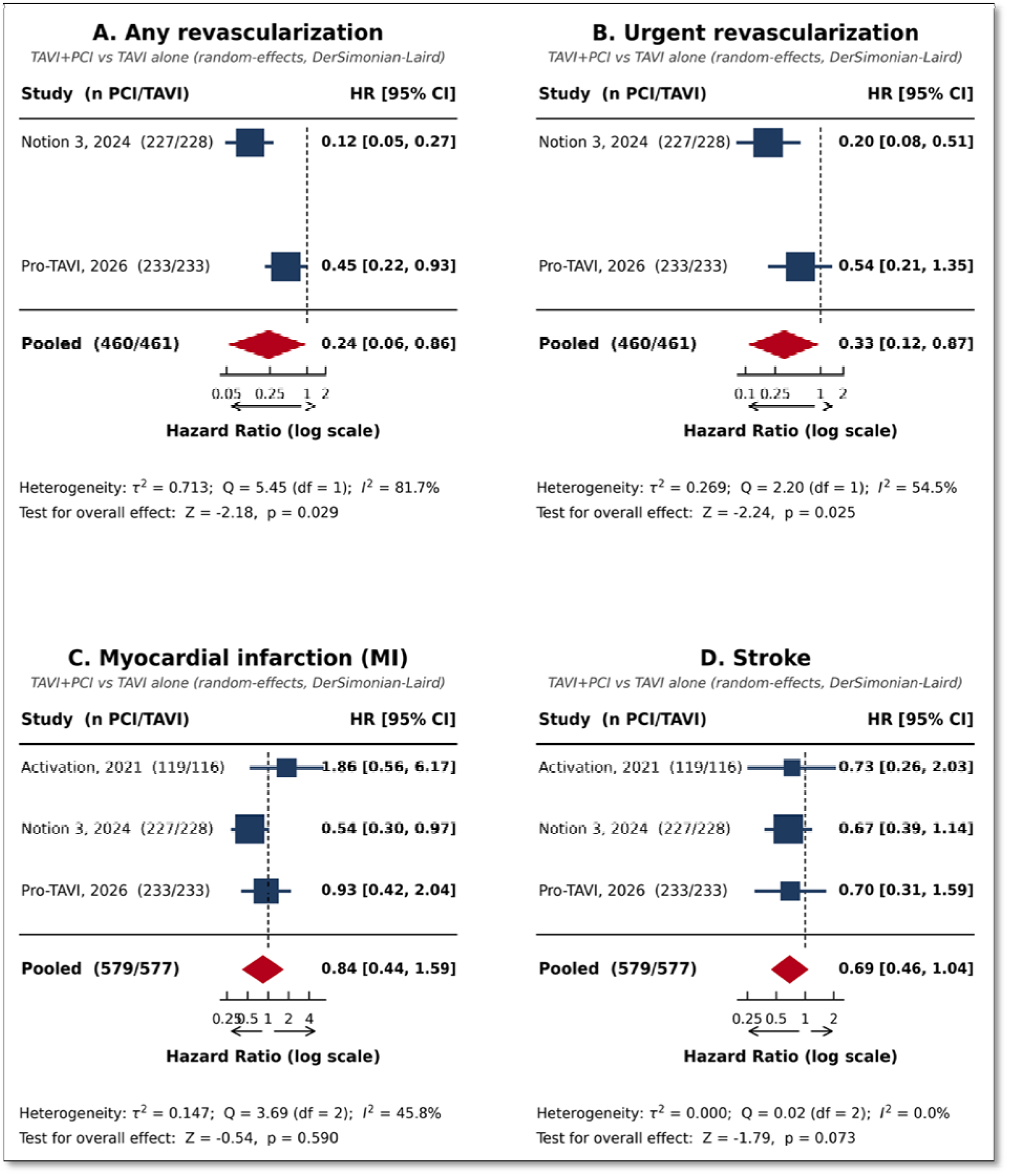
Forest Plots of Revascularization, MI, and Stroke Outcomes. This figure displays random-effects meta-analyses of secondary endpoints. Panel A: any revascularization (HR 0.24, 95% CI 0.06–0.86; p=0.029); Panel B: urgent revascularization (HR 0.33, 95% CI 0.12–0.87; p=0.025); Panel C: MI (HR 0.84, 95% CI 0.44–1.59; p=0.590); Panel D: stroke (HR 0.69, 95% CI 0.46–1.04; p=0.073). PCI significantly reduced revascularization endpoints, while MI was not significantly different and stroke showed a borderline trend favoring PCI. Blue squares represent individual trial HRs (size proportional to weight); horizontal lines denote 95% CIs. Red diamonds represent pooled estimates. The dashed vertical line marks HR=1.0. Abbreviations: CI, confidence interval; df, degrees of freedom; HR, hazard ratio; I², I-squared statistic for heterogeneity; MI, myocardial infarction; n, number of patients; PCI, percutaneous coronary intervention; Q, Cochran’s Q statistic; TAVI, transcatheter aortic valve implantation; τ², tau-squared; Z, test statistic for the overall effect.

### 3.8 Myocardial Infarction

The pooled effect on myocardial infarction (MI), reported across all three trials, favored TAVR plus PCI numerically but did not reach statistical significance (HR 0.84, 95% CI 0.44–1.59; p = 0.590), with moderate heterogeneity (I² = 45.8%, τ² = 0.147) **(Figure 3C).**

### 3.9 Stroke

Stroke trended in favor of TAVR plus PCI but did not achieve significance (HR 0.69, 95% CI 0.46–1.04; p = 0.073), with no detectable heterogeneity (I² = 0.0%) **(Figure 3D)**.

### 3.10 Bleeding outcomes

Bleeding endpoints significantly increased among patients receiving TAVR plus PCI. Any bleeding occurred almost twice as often in the PCI arm (pooled HR 1.96, 95% CI 1.28–3.00; p = 0.002), although heterogeneity across trials was substantial (I² = 71.4%, τ² = 0.101) and was driven by a higher event rate reported in the Pro-TAVI trial **(Figure 4A)**.

**Figure 4.**
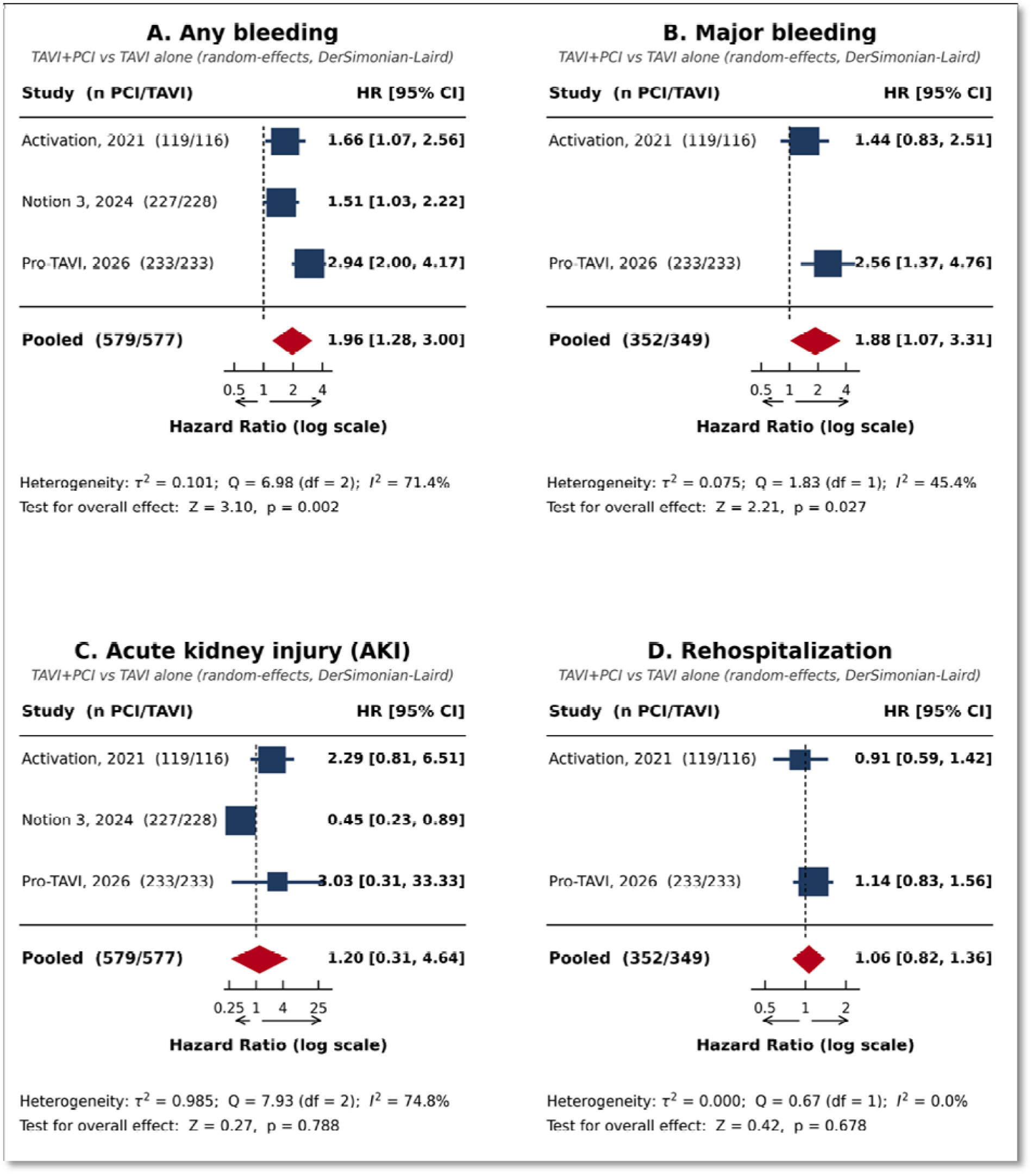
Forest Plots of Bleeding, AKI, and Rehospitalization Outcomes. This figure presents random-effects meta-analyses of safety endpoints. Panel A: any bleeding (HR 1.96, 95% CI 1.28–3.00; p=0.002; I²=71.4%); Panel B: major bleeding (HR 1.88, 95% CI 1.07–3.31; p=0.027; I²=45.4%); Panel C: AKI (HR 1.20, 95% CI 0.31–4.64; p=0.788; I²=74.8%); Panel D: rehospitalization (HR 1.06, 95% CI 0.82–1.36; p=0.678; I²=0.0%). PCI nearly doubled both any and major bleeding risk, while AKI and rehospitalization did not differ significantly. Blue squares represent individual trial HRs (size proportional to weight); horizontal lines denote 95% CIs. Red diamonds represent pooled estimates. The dashed vertical line marks HR=1.0. Abbreviations: AKI, acute kidney injury; CI, confidence interval; df, degrees of freedom; HR, hazard ratio; I², I-squared statistic for heterogeneity; n, number of patients; PCI, percutaneous coronary intervention; Q, Cochran’s Q statistic; TAVI, transcatheter aortic valve implantation; τ², tau-squared; Z, test statistic for the overall effect.

Major bleeding was reported by ACTIVATION and PRO-TAVI, while NOTION-3 reported event counts (26 vs 22) without a corresponding hazard ratio and was therefore excluded from this pooled estimate. Across the two contributing trials TAVR plus PCI was associated with a significantly increased risk of major bleeding (pooled HR 1.88, 95% CI 1.07–3.31; p = 0.027), with moderate heterogeneity (I² = 45.4%, τ² = 0.075) **(Figure 4B)**.

### 3.11 Acute Kidney Injury

The pooled risk of acute kidney injury (AKI) did not differ significantly between groups (HR 1.20, 95% CI 0.31–4.64; p = 0.788), but the estimate was characterized by very high between-study heterogeneity (I² = 74.8%, τ² = 0.985) reflecting markedly discordant trial-level findings **(Figure 4C).**

### 3.12 Rehospitalization

Rehospitalization, reported by two trials (ACTIVATION and Pro-TAVI), did not differ between groups (pooled HR 1.06, 95% CI 0.82–1.36; p = 0.678; I² = 0.0%) **(Figure 4D).**

### 3.13 Sensitivity analysis

Leave-one-out analyses showed stable pooled estimates for all-cause mortality, stroke, and any bleeding, regardless of which trial was excluded. The AKI estimate was highly sensitive (HRs 0.83–2.40), suggesting the null finding reflects counterbalancing trial-level effects rather than true absence. MI also showed directional instability (HRs 0.66–1.15); excluding NOTION-3 yielded a more pronounced effect, indicating its substantial contribution to heterogeneity. Outcomes informed by only two trials (cardiovascular mortality, major bleeding, revascularization, rehospitalization) could not be analyzed.

## 4. DISCUSSION

This systematic review and meta-analysis of three randomized controlled trials enrolling 1,156 patients provides a comprehensive synthesis of randomized evidence to date on the role of PCI before TAVR in patients with concomitant severe aortic stenosis and coronary artery disease.

The principal finding is that routine PCI before TAVR does not significantly reduce all-cause mortality (HR 0.88, 95% CI 0.67–1.17; I² = 0%) or cardiovascular death (HR 0.77, 95% CI 0.49–1.19). While PCI was associated with significant reductions in urgent revascularization (HR 0.33, 95% CI 0.12–0.87) and a numerical trend toward reduced stroke (HR 0.69, 95% CI 0.46–1.04), these potential ischemic benefits were offset by substantially higher risk of any bleeding (HR 1.96, 95% CI 1.28–3.00) and major bleeding (HR 1.88, 95% CI 1.07–3.31). These findings underscore a fundamental trade-off between a lower risk of ischemic events and a higher hemorrhagic risk that must be carefully weighed in clinical decision-making.

Across all three trials, routine PCI before TAVR did not reduce all-cause mortality, and the pooled estimate showed zero heterogeneity (I² = 0%). This finding aligns with the broader evidence about stable CAD, where landmark trials such as COURAGE and ISCHEMIA demonstrated that elective revascularization does not improve survival compared to medical treatment alone [12, 13]. Although the pathophysiology of myocardial ischemia in the setting of severe aortic stenosis differs fundamentally from isolated CAD; left ventricular hypertrophy, capillary rarefaction, and impaired coronary flow reserve create a supply–demand mismatch that could theoretically favor revascularization, this did not translate into a statistically significant mortality benefit with routine PCI. That being said, it’s important to point out that none of the three trials was powered to detect a difference in all-cause mortality as a standalone endpoint, and the pooled sample of 1,156 patients may still be insufficient to detect a modest but clinically meaningful mortality reduction; the confidence interval for the pooled estimate (0.67–1.17) does not exclude a potential mortality benefit of up to 33%, and the stability of this finding in leave-one-out sensitivity analyses (HR range 0.86–0.93), while reinforcing its robustness, underscores the need for a larger pooled dataset to definitively resolve this question.

The cardiovascular death analysis (HR 0.77, 95% CI 0.49–1.19) hints at a possible benefit of PCI, but the signal is too weak and the sample too small to draw firm conclusions. Only two trials contributed to this estimate, since hazard ratios for cardiovascular mortality were unavailable from ACTIVATION. NOTION-3 showed a more favorable trend (HR 0.67) than PRO-TAVI (HR 0.93), and the difference between them likely reflects, at least in part, the longer follow-up in NOTION-3 (2 years vs. 1 year) [10, 11]. If the benefit of revascularization on cardiovascular death takes time to emerge — as the late divergence of event curves in the ISCHEMIA trial would suggest — then 1-year follow-up may simply be too short to capture it [13].

The pooled effect on myocardial infarction (HR 0.84, 95% CI 0.44–1.59) was non-significant, with moderate heterogeneity (I² = 45.8%). This heterogeneity was driven primarily by the NOTION-3 trial, which reported a significant reduction in myocardial infarction with PCI (HR 0.54; 95% CI 0.30–0.97), in contrast to the ACTIVATION and PRO-TAVI trials, which showed no significant difference [9–11]. The leave-one-out analysis confirmed that exclusion of NOTION-3 shifted the pooled estimate toward the null (HR 1.15). This is most likely attributed to the fact that NOTION-3 employed more accurate assessment to define significant CAD; FFR ≤ 0.80, or severe angiographic stenosis (≥ 90%), whereas ACTIVATION used a purely anatomic threshold (≥ 70% diameter stenosis). Also, FFR-guided PCI has been shown to improve outcomes compared with angiography-guided PCI in the broader stable CAD population, and the use of physiologic assessment in NOTION-3 may have selected a population more likely to benefit from revascularization [9, 10]. Additionally, the longer follow-up in NOTION-3 may have allowed more time for the protective effect against spontaneous MI to manifest [10].

The pooled estimate for stroke (HR 0.69, 95% CI 0.46–1.04; p = 0.073) showed a borderline significant trend favoring PCI, with no heterogeneity (I² = 0%). This finding was stable in leave-one-out sensitivity analyses (HR range 0.68–0.71), suggesting a consistent direction of effect across trials. The mechanism, however, is not obvious, as PCI itself carries a risk of periprocedural cardioembolic stroke. One possible explanation is that by addressing hemodynamically significant coronary lesions before TAVR, PCI may improve hemodynamic stability during the valve implantation procedure, thereby reducing the risk of hypotension-related cerebral hypoperfusion. Alternatively, the reduction in subsequent urgent revascularization procedures — each of which carries its own stroke risk — may contribute to the lower stroke incidence in the PCI group. Another possible explanation is that the addition of DAPT might have prevented non-cardioembolic stroke. Although DAPT is not recommended for primary prevention of stroke, according to the **PEGASUS-TIMI 54,** adding ticagrelor to patients with prior MI, reduced the ischemic events at 3-year follow-up compared to placebo [14]. Either way, this finding needs confirmation in larger studies before it can influence practice.

If there is one finding in this meta-analysis that should change how clinicians think about routine PCI before TAVR, it is the bleeding signal. Both any bleeding (HR 1.96, 95% CI 1.28–3.00) and major bleeding (HR 1.88, 95% CI 1.07–3.31) were significantly increased with PCI, and every individual trial pointed in the same direction. The leave-one-out analyses confirmed the robustness of this result. The POPular TAVI trial showed that adding clopidogrel to aspirin after TAVR increases bleeding risk by 76% without reducing ischemic events [15]. A subsequent meta-analysis of SAPT versus DAPT after TAVR corroborated this, reporting a 3-fold increase in major bleeding with DAPT and no ischemic benefit [16].

The clinical significance of these findings should not be underestimated. Major bleeding following TAVR is not a benign complication, particularly in a population that is predominantly elderly, often frail, and burdened with comorbidities that amplify hemorrhagic risk. Notably, substantial heterogeneity was observed for any bleeding (I² = 71.4%), driven largely by higher event rates in PRO-TAVI. This variability is likely attributable to differences in bleeding definitions, antithrombotic regimens, and baseline patient characteristics. Despite this heterogeneity, the direction of effect remained consistent across studies, reinforcing the robustness of the overall findings.

PCI before TAVR was associated with marked reductions in both any revascularization (HR 0.24, 95% CI 0.06–0.86) and urgent revascularization (HR 0.33, 95% CI 0.12–0.87). The high heterogeneity for any revascularization (I² = 81.7%), however, reflects a considerably larger effect size in NOTION-3 than in PRO-TAVI, which may be related to potential ascertainment or surveillance bias. Because these were open-label trials, clinicians caring for patients in the conservative arm knew that obstructive coronary disease had been left untreated and may have lowered their threshold for invasive evaluation when chest pain or dyspnea developed after TAVR. The NOTION-3 investigators themselves raised this possibility, noting that awareness of an unrevascularized lesion may have prompted more aggressive workup of ambiguous symptoms in the conservative group. However, a 76% relative risk reduction is too large to be attributed solely to surveillance bias, and the consistent direction of effect across trials supports a genuine underlying treatment effect.

The reduction in urgent revascularization is clinically meaningful, since these procedures after TAVR can be risky and technically challenging. Urgent revascularization carries higher mortality in elderly TAVR patients compared to the general population [17]. Also, coronary cannulation fails in up to 10% of post-TAVR PCI attempts, and access difficulty is more prominent with supra-annular self-expanding valve [18, 19]. Approximately 60% of NOTION-3 patients received self-expanding valves, meaning that subsequent PCI in the conservative arm was likely more technically demanding, required more contrast, and carried higher procedural risk than PCI in a non-TAVR patient. This does not invalidate the trial finding, but it does highlight that for patients with untreated obstructive lesions who will receive a self-expanding valve, deferring PCI carries a future procedural cost separate from the ischemic risk itself.

The pooled estimate for acute kidney injury (HR 1.20, 95% CI 0.31–4.64) was non-significant but characterized by very high heterogeneity (I² = 74.8%). The inconsistency across trials may reflect differences in contrast volumes, staging strategies (same day vs. staged PCI), baseline renal function, and AKI definitions. Given the vulnerability of the elderly TAVR population to contrast-induced nephropathy, the potential for increased AKI with PCI warrants continued surveillance in future studies.

Rehospitalization rates did not differ between groups (HR 1.06, 95% CI 0.82–1.36; I² = 0%), based on data from ACTIVATION and PRO-TAVI. This finding suggests that PCI before TAVR neither increases nor decreases the overall burden of hospitalization. The ACTIVATION trial used rehospitalization as a component of its primary composite endpoint and found no difference between arms. The absence of a signal for rehospitalization is consistent with the overall neutral effect of PCI on mortality and the offsetting effects on ischemic and bleeding endpoints.

## 5. Limitations

This meta-analysis has several important limitations. First, only three RCTs were available, limiting statistical power. Second, follow-up durations differed: two trials reported 1-year outcomes, NOTION-3 reported 2-year, so hazard ratios were used. Third, primary composite endpoints had varying definitions, precluding pooled analysis. All three trials were open-label, introducing potential performance and detection bias. ACTIVATION was terminated early due to slow enrolment, potentially affecting estimate stability. Finally, subgroup analyses by CAD severity (e.g., SYNTAX score) were unfeasible without individual patient data.

## 6. CONCLUSIONS

In patients with severe aortic stenosis and concomitant coronary artery disease undergoing TAVR, this meta-analysis of three randomized controlled trials demonstrates that routine PCI before TAVR does not reduce all-cause mortality or cardiovascular death. While PCI is associated with significant reductions in urgent revascularization and a trend toward reduced stroke, these potential ischemic benefits are counterbalanced by a significant 88–96% increase in bleeding events. These findings support a selective rather than routine PCI strategy before TAVR, with decisions individualized based on coronary anatomy and physiologic significance, symptom burden, bleeding risk, and frailty status.

## Clinical Impact

This meta-analysis carries important clinical implications. First, the absence of mortality benefit argues against routine PCI in patients with concomitant CAD, consistent with the 2020 ACC/AHA Class 2a recommendation favoring PCI in LM, proximal LAD and bifurcation CAD [1].

Second, the reduction in urgent revascularization and stroke trend suggests selected patients may derive ischemic benefit. NOTION-3, using FFR-guided or severe stenosis criteria, showed the most favorable results, with subgroup analysis confirming benefit only in non-frail patients [20]. Pro-TAVI failed non-inferiority for its death/MI/stroke endpoint (HR 4.3; 95% CI −2.7 to 11.4) [11].

Third, the higher bleeding highlights the need for shorter DAPT and contemporary antithrombotic strategies, including single antiplatelet therapy post-TAVR per POPular TAVI [15, 21, 22].

## Future Perspectives

Several ongoing trials should help clarify remaining uncertainties. The TAVI PCI trial (986 planned patients) is investigating the optimal sequencing of PCI and TAVR (before vs after), and the COMPLETE TAVR trial (NCT04634240) is evaluating staged complete revascularization after TAVR versus medical management alone. Future studies incorporating physiologic assessment of coronary lesions (e.g., FFR-guided PCI) and longer follow-up periods will be critical to refining treatment algorithms.

## Data Availability

All data produced in the present study are available upon reasonable request to the authors

## Abbreviation List

AKI: acute kidney injury
AS: aortic stenosis
CAD: coronary artery disease
CI: confidence interval
DAPT: dual antiplatelet therapy
FFR: fractional flow reserve
HR: hazard ratio
I²: heterogeneity statistic
MACE: major adverse cardiovascular events
MeSH: Medical Subject Headings
MI: myocardial infarction
PCI: percutaneous coronary intervention
PRISMA: Preferred Reporting Items for Systematic Reviews and Meta-Analyses
RCT: randomized controlled trial
RFR: resting full-cycle ratio
RoB-2: Cochrane Risk of Bias 2
SAPT: single antiplatelet therapy
STS: Society of Thoracic Surgeons
SYNTAX: Synergy Between Percutaneous Coronary Intervention with Taxus and Cardiac Surgery
TAVI: transcatheter aortic valve implantation
TAVR: transcatheter aortic valve replacement
VARC-3: Valve Academic Research Consortium-3 τ**²:** between-study variance

## Acknowledgment

The authors wish to express their sincere gratitude to Dr. John Mandrola for his dedication to medical education and his thoughtful, evidence-based commentary on contemporary cardiovascular literature through his podcast *This Week in Cardiology*. His work has consistently fostered critical appraisal of the evidence and inspired a more rigorous, skeptical approach to interpreting clinical trial data — a perspective that has meaningfully shaped our analytical framework [23].

## Artificial intelligence disclosure

Claude AI was used to assist with the design of the central illustration and to improve grammar and language clarity in the manuscript. The authors reviewed and edited all AI-assisted content and take full responsibility for the accuracy, integrity, and final content of the manuscript.

## Authors’ Contributions

Dina Soliman: concept/design, data collection, data analysis/interpretation, drafting article, critical revision of article.

John Abdelmalek: concept/design, data collection, data analysis/interpretation, critical revision of article.

Chanokporn Puchongmart: data collection, data analysis/interpretation

Tulaton Sodsri: Statistical analysis

Nithila Sivakumar: data collection, drafting article

Zhaunn Sly: approval of article.

